# Dual-mode near-infrared multispectral imaging system equipped with deep learning models improves the identification of cancer foci in breast cancer specimens

**DOI:** 10.1101/2022.10.27.22281556

**Authors:** Jun Liao, Lingling Zhang, Han Wang, Ziqi Bai, Meng Zhang, Dandan Han, Zhanli Jia, Yao Liu, Chenchen Qin, ShuYao Niu, Hong Bu, Jianhua Yao, Yueping Liu

## Abstract

For surgically resected breast cancer samples, it is challenging to perform specimen sampling by visual inspection, especially when the tumor bed shrinks after neoadjuvant therapy in breast cancer. In this study, we developed a dual-mode near-infrared multispectral imaging system (DNMIS) to overcome the human visual perceptual limitations and obtain richer sample tissue information by acquiring reflection and transmission images covering visible to NIR-II spectrum range (400–1700 nm). Additionally, we used artificial intelligence (AI) for segmentation of the rich multispectral data. We compared DNMIS with the conventional sampling methods, regular visual inspection and a cabinet X-ray imaging system, using data from 80 breast cancer specimens. DNMIS demonstrated better tissue contrast and eliminated the interference of surgical inks on the breast tissue surface, helping pathologists find the tumor area which is easy to be overlooked with visual inspection. Statistically, AI-powered DNMIS provided a higher tumor sensitivity (95.9% vs visual inspection 88.4% and X-rays 92.8%), especially for breast samples after neoadjuvant therapy (90.3% vs visual inspection 68.6% and X-rays 81.8%). We infer that DNMIS can improve the breast tumor specimen sampling work by helping pathologists avoid missing out tumor foci.

## Introduction

Nowadays, a large number of clinical studies jointly conducted by artificial intelligence (AI) engineers and pathologists focus on the interpretation of biomarkers and pathological microscope image analysis, while the identification and sampling of lesions on gross specimens are neglected. Pathology is the gold standard for diagnosis, and incomplete sampling has a great impact on the accuracy of lesion identification and subsequent accurate diagnosis. Traditional pathological sampling^1-3^ mainly relies on the visual inspection of pathologists by color and texture differences. However, the doctor’s experience is subjective, and the human eye has a limited range of perception (visible light range 400–700 nm). In addition, in the field of breast cancer sampling, the evaluation of intraoperative surgical margins for breast conserving surgery and evaluation of tumor beds after neoadjuvant therapy for breast cancer have always been challenging for pathologists. Especially in patients who received neoadjuvant therapy, the residual tumor is scattered non-centripetally, which makes it extremely difficult to identify the boundary by visual inspection during the sampling process.

The cabinet X-ray imaging system can assist the pathological sampling of postoperative breast samples to a certain extent^4,5^. However, owing to the large difference in the density between the breast cancer foci and adipose tissue, the density-based transmission imaging method of X-rays can distinguish adequately between the tumor tissue and fat, but has a weaker discrimination ability for fibrous tissue and tumor tissue with similar densities. Some techniques for intraoperative detection of tumor margins may be useful to assist postoperative specimen sampling, such as ultrasound^6-8^, optical coherence tomography (OCT)^9,10^, photoacoustic imaging^11-12^, computed tomography (CT)^13^, terahertz (THz)^14-21^, Raman spectroscopy^22-24^, and hyperspectral imaging^25-30^. However, these methods require special technicians to operate the equipment, and have disadvantages such as a long acquisition time, low image resolution, potential radiation, and high cost.

In recent years, near infrared region two (NIR-II, 1000–1700 nm) has gained considerable attention because of its better imaging quality and penetration depth in living organisms^31,32^ than the visible bands. The advantages of NIR-II, such as low scattering in living organisms, large penetration depth, non-destructive and non-contact imaging, and low training requirements for operators, have attracted widespread attention in the field of life sciences and clinical surgery^33^. This is the first study to propose a dual-mode near-infrared multispectral assisted sampling system (DNMIS) covering the visible and NIR window, which is label-free compared to NIR-II fluorescence imaging. Although we claimed “near infrared” in DNMIS, we still added three visible bands (red: 625 nm, green: 525 nm, blue: 465 nm) to DNMIS to produce a color image for comparing and verifying if the visible band has a positive contribution when identifying a tumor in a specimen. The DNMIS contains a total of 13 narrow-band spectral images covering visible-NIR-II in the range of 400–1700 nm. Both reflection and transmission imaging modes are included. Cross-polarization is introduced into reflection imaging to eliminate glare from the sample surface. From the collected data, we synthesized pseudo-color images with rich information for direct reference by doctors. In addition, we also used a deep learning algorithm to analyze the entire multispectral images to achieve pixelwise tissue type segmentation. Compared to conventional color images, the multispectral images captured by DNMIS have better tissue contrast, clearer tissue boundaries, and can even visualize tissue through surgical dye inks to help doctors to accurately draw materials. Quantitative research has shown that when using deep learning to segment tissue multispectral data, the sensitivity of identifying tumor tissue in breast cancer samples is 95.9%, which is higher than that using visual inspection (88.4%) and X-rays (92.8%), especially for breast samples after neoadjuvant therapy. DNMIS sensitivity was 90.3% compared to 68.6% for visual inspection and 81.8% for X-rays. DNMIS can greatly improve the accuracy and confidence of doctors, thereby ensuring the lesion integrity of subsequent pathological analysis.

## Results

### Scheme of the proposed DNMIS system

As shown in Fig.1a, pathological specimen sampling is an important part of the routine work of pathologists. To improve the confidence of pathologists while sampling, we developed the DNMIS as shown in Fig. 1b. The DNMIS consists of three main components: broadband sensing (400–1700 nm) camera, autofocus lens, and multispectral light source (400–1700 nm). As shown in Fig. 1b, the specimen is placed on a translucent frosted acrylic sheet. Each multispectral image acquisition collects 13 images, including nine reflected and four transmitted images. Two obliquely illuminated light sources positioned above the sample provide reflected illumination on the sample. The two reflection light sources are identical, each containing the following nine types of narrow-band LEDs: three visible bands: 465 nm (Blue), 525 nm (Green), and 625 nm (Red) and six NIR-II bands: 1050 nm, 1200 nm, 1300 nm, 1450 nm, 1550 nm, and 1650 nm. A backlight located below the sample provides transmitted illumination for the sample, and contains four near-infrared wavelengths: 760 nm, 850 nm, 1050 nm, and 1300 nm. Fig.1c shows the spectral shape of these LEDs. We selected these multispectral bands based on a combination of previous hyperspectral pre-experiments (Supplementary video) and the commercial availability of NIR-II LEDs. For example, the four selected transmission bands have a strong transmission ability in the breast tissue, while the selected wavelength of NIR-II can reflect the higher contrast between different breast tissues. The two NIR-I wavelengths (760nm and 850nm) are selected only as transmitted lights instead of reflection lights since they have high tissue transmission in transmitted imaging but provide poor tissue contrast in reflected imaging as shown in the supplementary video. The three narrow-band visible wavelengths we selected can be used for comparison with the NIR images as well as for verifying if the visible bands have a positive contribution in tumor segmentation. Please check the detailed description of the hardware components in the methods section.

**Fig. 1.**
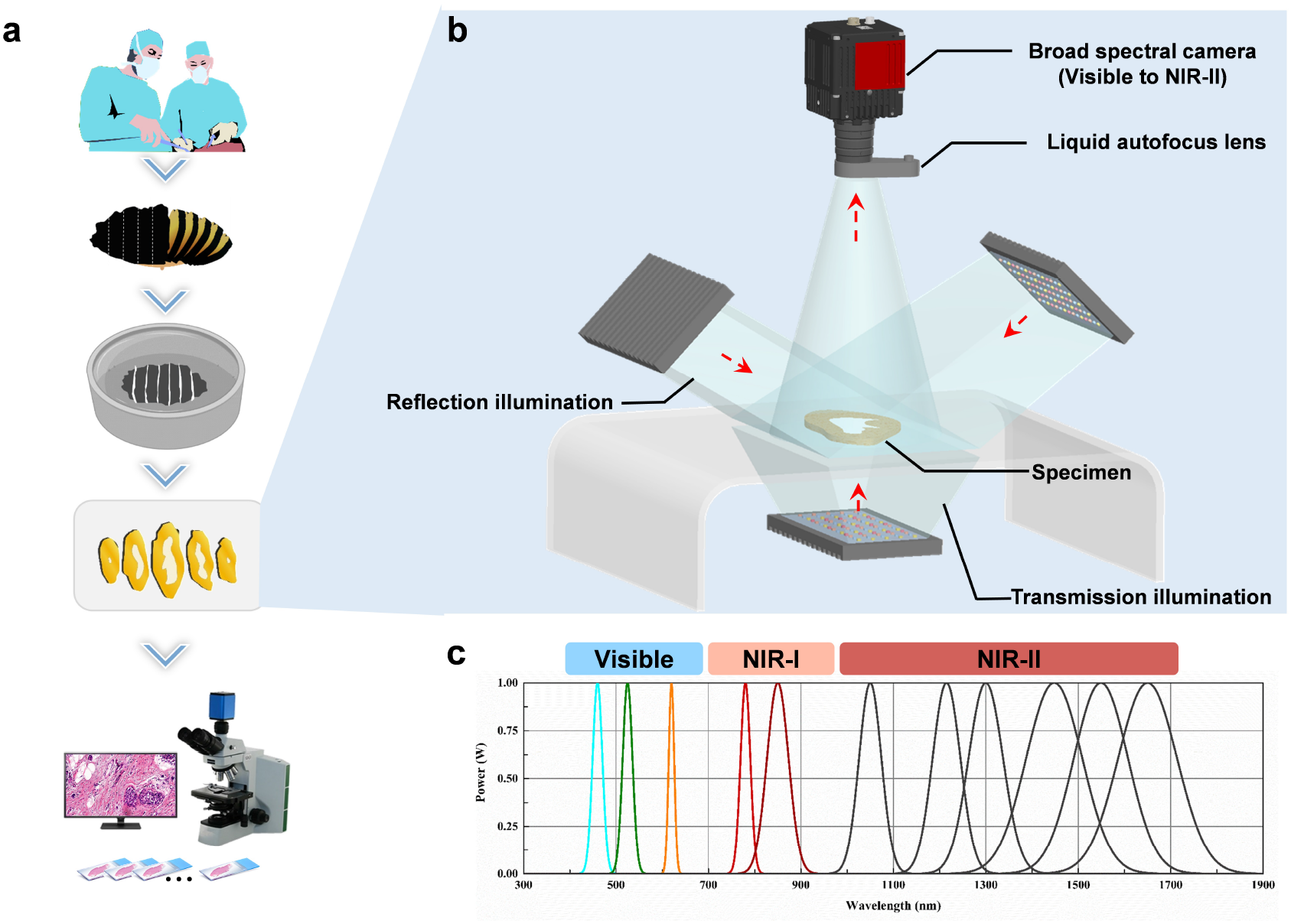
The proposed NIR-II multispectral imaging system for pathological specimen sampling. a. The surgical specimens were cut and fixed, and tissue pieces were cut for use. After all images were collected, the specimens were grossly collected, routinely dehydrated, made into pathology slides and observed under a microscope. b. We introduced DNMIS to assist specimen sampling. The main hardware components of DNMIS contains: multispectral light sources, a broad spectral sensing camera(400-1700nm) and an autofocus lens set. c. The LED spectral profiles employed in DNMIS.

### Statistical comparison of our work and conventional specimen sampling methods

Although the multispectral information of samples collected by DNMIS is rich, it is difficult for pathologists to read up to 13 images simultaneously and make decisions quickly. AI, the data-driven approach, is best used to find subtle features within massive amounts of information and achieve quick results. Therefore, we used the U-net, a deep learning network, to analyze the 3D multispectral information (2D spatial + 1D spectral) and help pathologists in segmenting the boundaries of different tissues. Fig. 2a shows the details of the multispectral data and other types of imaging data that were collected. The regular color image was captured using a Canon digital camera to simulate visual inspection. The X-ray image was collected by the cabinet X-rays imaging system BioVision (Faxitron Bioptics, LLC, IL, USA). We also developed a whole slide images (WSI) stitching software to stitch the scanned WSI fragments to recover the digital large slide of the sample where a breast lesion needs several slides to cover its entire area. As microscopic images are the gold standard for cancer diagnosis, we obtained the ground truth of tissue labeling after registering the stitched WSI with the gross images.

**Fig.2.**
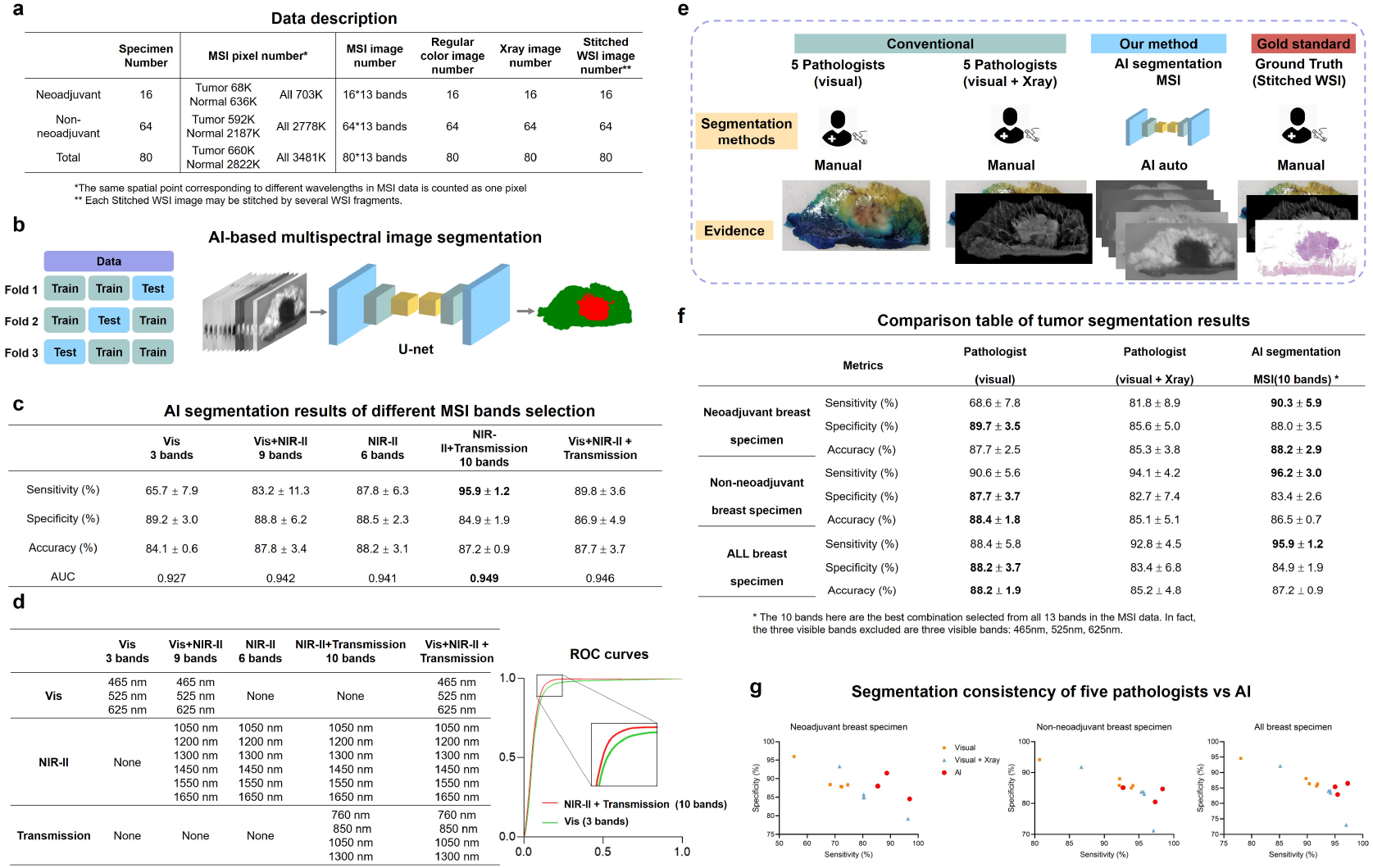
AI-assisted DNMIS segmentation performance vs conventional methods. a. data description table of all the collected data. b. We use U-net and three-fold cross-validation to train and test the DNMIS data. c. AI segmentation results of different MSI bands selection. d. The table shows the waveband combination details. The ROC curve shows the visible only (the worst combination) and NIR-II + transmission (the best combination) in c. Vis denotes visible bands. e. the scheme of tumor segmentation comparison experiments between AI-assisted DNMIS and conventional methods. The segmentation ground truth is determined using the stitched WSI as primary evidence. f. the comparison table of tumor segmentation results. g. Sensitivity and specificity in the segmentation consistency of different specimens by five pathologists and AI.

We used U-net and 3-fold cross validation methods to train and test the tumor segmentation performance of DNMIS (Fig. 2b). We tested combinations of four multispectral bands to explore whether visible, NIR-II, and transmitted light positively contribute to the final result. The table in Fig. 2c shows that the three visible bands had a negative contribution, while NIR-II reflected and transmitted lights had a positive contribution to the segmentation ability. Since for the task of specimen sampling, avoid missing tumor foci (high sensitivity) is more important than less trivial work (high specificity), we regarded the NIR-II + transmission (10 bands) option as our best result as it had the highest sensitivity (95.9%) for tumor identification and competitive overall segmentation ability (Accuracy: 87.2%, area under the curve [AUC]: 0.949) compared to other combinations. Fig. 2d shows the detailed waveband combinations and receiver operating characteristic (ROC) curve shows the visible only (the worst combination) and NIR-II + transmission (the best combination) in Fig. 2c.

To compare with the conventional sampling methods, we recruited five pathologists with more than 3 years of clinical experience to do three sets of comparative experiments. As shown in Fig. 2e, the five pathologists manually sketched the tumor margin using the regular color image as the evidence. After 2 weeks as the forgetting period, they were asked to repeat the sketch using both the regular color image and X-ray image as the evidence. For our proposed method, tumor segmentation in the manual sketch is not required. In the end, the five pathologists and one senior pathologist (with over 10 years of professional experience) jointly specified the ground truth of the segmentation results according to the stitched WSI (primary evidence) and other imaging evidence.

Fig. 2f shows the comparison table of the above-mentioned experiments. It can be seen that for neoadjuvant breast specimens, AI segmentation provides outstanding sensitivity and competitive specificity, which means DNMIS is less likely to miss tumor positive areas. For non-neoadjuvant breast specimens, the high sensitivity advantage of DNMIS still exists. For the task of specimen sampling, avoiding missing tumor area (high sensitivity) is more important than higher recognition ability of negative samples (high specificity).

Furthermore, manual sketching is subjective and the sketch consistency of the tumor area among the five pathologists was poor, as shown in Fig. 2g. The standard deviation of the five pathologists of marking tumor area is shown in Fig. 2f. Different from manual annotation, the trained AI network can get exactly the same result every time it predicts the same input data. The AI based method in Fig. 2g has three points since we use 3-fold testing strategy as shown in Fig. 2b.

### NIR-II imaging shows clear borders in postoperative breast cancer specimens after non-neoadjuvant therapy

During the process of pathological sampling, a clear tumor boundary is helpful for pathologists to judge the extent of the tumor and accurately measure the maximum diameter of the tumor. While evaluating the surgical margins of the specimen during breast-conserving surgery, a clear tumor boundary can help the doctors to observe the distance between the lesion and the surgical margin, thereby avoiding positive postoperative margins due to missed detection and reduce the reoperation rate. In the present study, experimental results showed that NIR-II imaging has a good discriminative ability for breast cancer tumor bed. The spectral curves of the cancer tissue, adipose tissue, and normal breast tissue with different characteristics are shown in Fig. 3f, demonstrating the different colors discernible by visual inspection in the synthetic pseudo-color image, in addition to revealing clear tumor boundaries (Fig.3b1-3).

**Fig 3.**
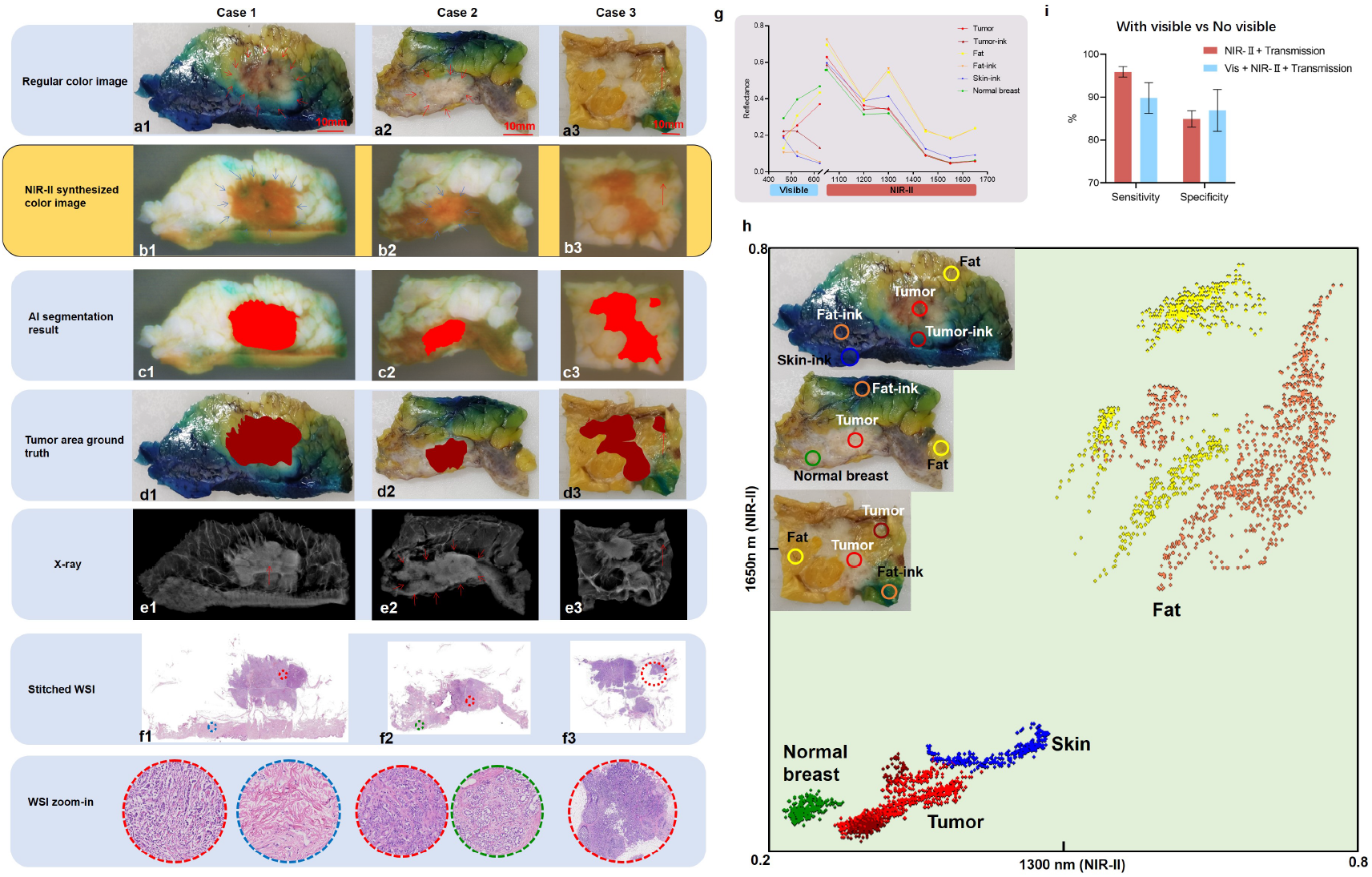
The performance of NIR-II imaging in different breast cancer samples. Case 1: (a1) Regular color image, the red arrows indicate the tumor area, the tumor boundaries are unclear by visual inspection. (b1) NIR-II synthesized color image, the tumor tissue is clearly demarcated (blue arrows). (c1): AI segmentation Result. (d1): The real tumor range marked with reference to the gold standard. (e1): In this case, the X-ray also showed good results (tumor: red arrow). (f1): Stitched WSI image, red circle shows invasive carcinomas, blue circle shows normal tissues. Case 2: (a2) In this case, the visible tumor border may be the area surrounded by red arrow. (b2): Invasive cancer appears brighter than surrounding tissue in NIR-II synthesized color image. (c2): AI segmentation result. The tumor region is correctly identified. (e2): The tumor area shown by X-ray is obviously larger than the real tumor bed shown by d2. (f2): Under the microscope, the red circle refers to invasive breast cancer, the area indicated by the blue circle shows adenosis. Case 3: (a3) In the regular color image, suspicious cancer foci are found (pointed by red arrow). (b3) NIR-II synthesized color image shows the lesion (see red arrow). (c3): AI segmentation identifies tumor at this site. (e3): No information provided in the X-ray. (f3): WSI confirmed here (shown in red circle) as invasive cancer tissue. (g): Spectral curves of different tissues. (h): Scatter plots projected by 1300nm and 1650nm DNMIS reflected image. (i): Sensitivity and specificity performance difference of AI-assisted DNMIS incorporating visible range bands or not.

At the same time, NIR-II imaging can also eliminate the influence of different color inks (Fig. 3b1). As the curve shows in Fig. 3f, the ink-stained adipose tissue and conventional adipose tissue have no regularity in the visible range, but are very close to the NIR-II spectral curve, which means that the ink is transparent in the NIR-II region, so that it cannot interfere with the imaging of fat. We compared the segmentation results of incorporating the visible range bands or not to determine whether the visible range bands have a negative contribution to the segmentation process. The statistics in Fig. 3h confirm this point.

As shown in Fig. 3 (Case 2), the breast cancer tissue in the NIR-II synthesized color image shows a brighter color than the surrounding adenosis, which is easier to identify than the regular color image (Fig. 3b2), and can intuitively reflect the relationship between the tumor and resection margins. In Fig. 3b1-b3, the NIR-II synthesized color image was synthesized as follows: 1050 nm was put into the red channel, 1300 nm into the green channel, and 1650 nm into the blue channel. NIR-II imaging can also help pathologists to identify suspicious lesions. As shown in Fig.3(Case 3), the area indicated by the red arrow in the regular color image has suspicious lesions (Fig. 3 a3), The NIR-II synthesized color image shows the same color features as the tumor tissue (Fig.3 b3), and AI segmented images are identified as tumor tissue (Fig.3 b3). It was confirmed that this is an invasive carcinoma in WSI (Fig. 3f3). The X-ray image does not give more hints (Fig.3 e3).

In Fig. 3g, we can observe that 1300 nm and 1650 nm provide good contrast between different tissues. Therefore, in Fig. 3h, we used two NIR-II wavebands (1300 nm and 1650 nm) to project all the circled area (top left corner in Fig. 3h) by the reflectance intensity. We can determine the clustering of the same type of tissue, with or without ink coverage. Here we obtained good tissue discrimination for this sample using only two wavelength projections. However, the tissue spectrum of different samples may have a small range of deviation, so the more wavelengths involved in the segmentation, the more accurate and robust the segmentation will be, although more wavelengths will bring redundant information. Statistically, among all the 80 breast specimens, adding visible bands to the AI input lowered the sensitivity of tumor identification, as shown in Fig. 3i.

### NIR-II synthesized color image shows human-unrecognizable tumor tissues in the tumor bed after neoadjuvant therapy

At present, the Residual Cancer Burden (RCB) score recommended by the Breast International Group-North American Breast Cancer Group (BIG-NABCG) needs to include factors such as the largest tumor diameter, proportion of in situ carcinoma, and proportion of invasive cancer for pathological evaluation of breast cancer patients after neoadjuvant therapy ^1,34^. Therefore, accurate data can only be obtained by a comprehensive and complete general sampling. After neoadjuvant therapy, the tumor bed of breast cancer shows different regression patterns. Especially when the tumor shows a non-centripetal regression pattern, it is difficult to distinguish the scattered tumor tissue in the tumor bed with visual inspection. Fig. 4a1-h1 shows a case with concentric regression of the tumor bed after neoadjuvant therapy. In the regular color image, the border of the tumor is unclear and irregular (Fig. 4 a1). In the NIR-II synthesized color image, the area surrounded by blue arrows is the location of the tumor bed (Fig.4 c1, d1). The AI segmentation result also accurately predicted the extent of the tumor bed (Fig.4 g1).

**Fig.4.**
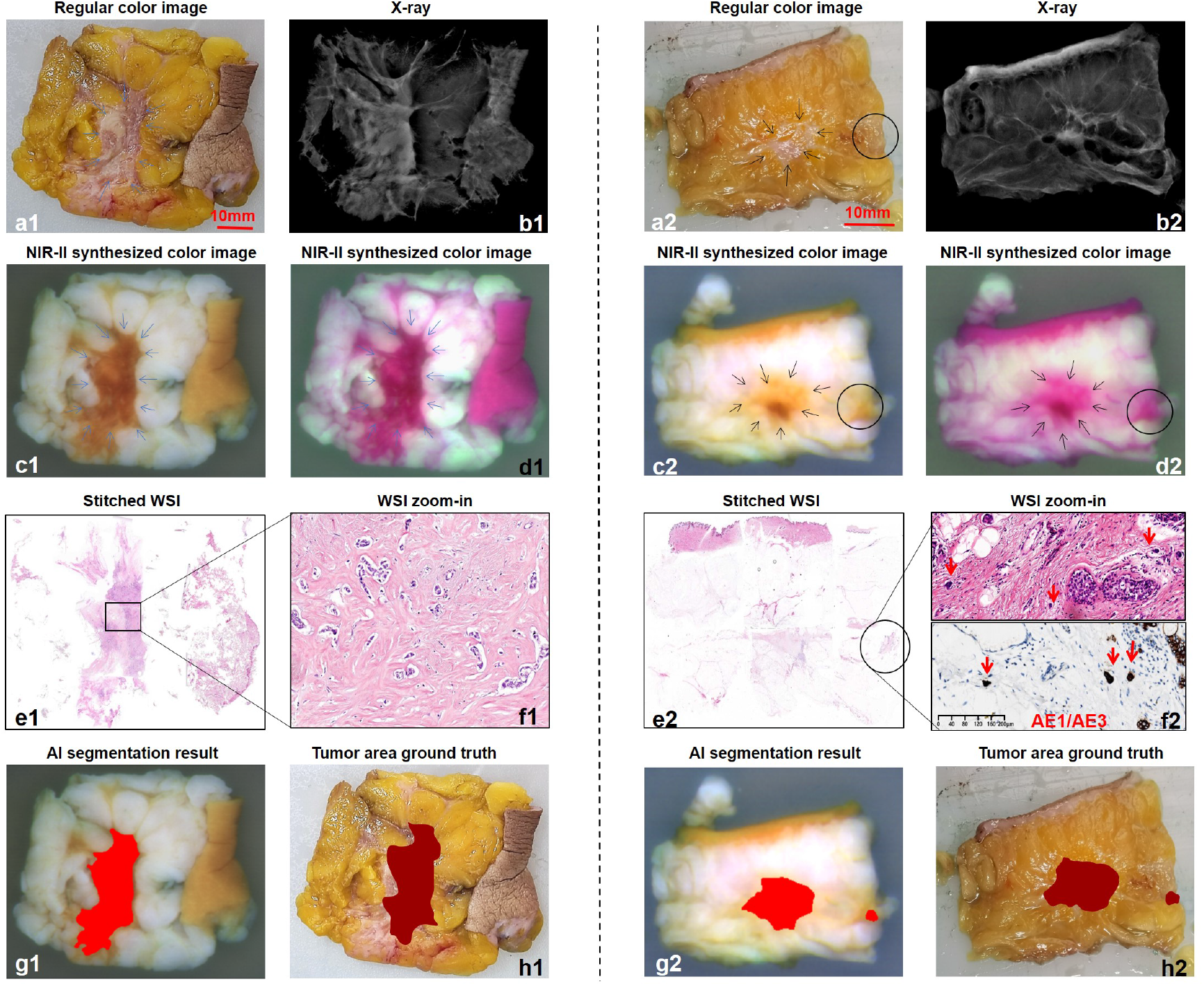
DNMIS improves the sampling of breast cancer samples after neoadjuvant therapy. (a1, a2): Regular color image. Suspicious lesions (black circle in a2). (b1, b2): X-ray image. (c1, c2): NIR-II synthesized color image, the synthesis method is R: 1050nm, G: 1300nm, B: 1650nm. and the area surrounded by blue arrows indicates a fibrotic tumor bed. (d1, d2): NIR-II synthesized color image, the synthetic method is: R: 1050nm; G: 1650nm; B: 1300nm. (e1, e2): Stitched WSI. (f1): Microscopic representation of the lesion in the black box magnified. (f2): Enlarged image in the black circle of e2, the red arrows point to the degenerated tumor cells. Immunohistochemical staining of CK showed strong positive expression in degenerated tumor cells. (g1-2): AI segmentation Result. (h1-2): Tumor area ground truth.

The results of this experiment show that NIR-II imaging has certain penetration capability, and can detect deeper lesions and identify tumor tissues that cannot be distinguished by visual inspection. As shown in Fig. 4a2-h2, in a specimen after neoadjuvant therapy with a non-centripetal retraction pattern, a2 shows the section of the tumor as seen by visual inspection. In the NIR-II synthesized color image (Fig. 4 c2 and d2), the suspected fibrotic tumor bed is shown by a black circle. The degenerated tumor cells (Fig.4f2) indicated by the red arrow in this area were confirmed to be tumor cells by immunohistochemical AE1/AE3 staining. This area is not obvious in the regular color image (Fig.4 a2) and X-ray image (Fig.4 b2), whereas AI segmentation accurately predicted the tumor tissue (Fig.4 g2).

### Infrared transmitted light further improves the identification of foci in multispectral images

Breast cancer foci are irregular in three-dimensions. The tumor tissue information displayed by the cut plane is only a small part on the surface. In order to obtain deeper tumor bed information, we also added image acquisition of near-infrared transmitted light in DNMIS. In rare cases, normal dense breast tissue may exhibit a similar image to tumor tissue (Fig. 5c, indicated by the green circle), which corresponds to normal breast tissue in WSI (Fig. 5 e, f). In transmitted light at 1300 nm (Fig. 5d), the light transmittance of tumor tissue is significantly worse than that of normal tissue, and the transmitted light image can assist in distinguishing between tumor and non-tumor tissues. AI segmentation results also made accurate predictions (Fig. 5g). The section intensity curve in Fig. i1 and i2 shows that the circled normal area (green dash line) has intensity closer to that of the tumor area (red dash line), making it prone to classify this normal area as false positive. However, in Fig. j1 and j2, the intensity of the circled area (green dash line) shows a good contrast with the tumor area (red dash line) in transmitted light at 1300 nm. Note that the X-ray (Fig. 5b) image is intensity reversed, which makes the tumor area brighter than the fat area (Fig. 5i2), while in the transmitted image, the tumor area is darker than the fat area (Fig. 5j2). The background intensity difference also proves this point.

**Fig. 5.**
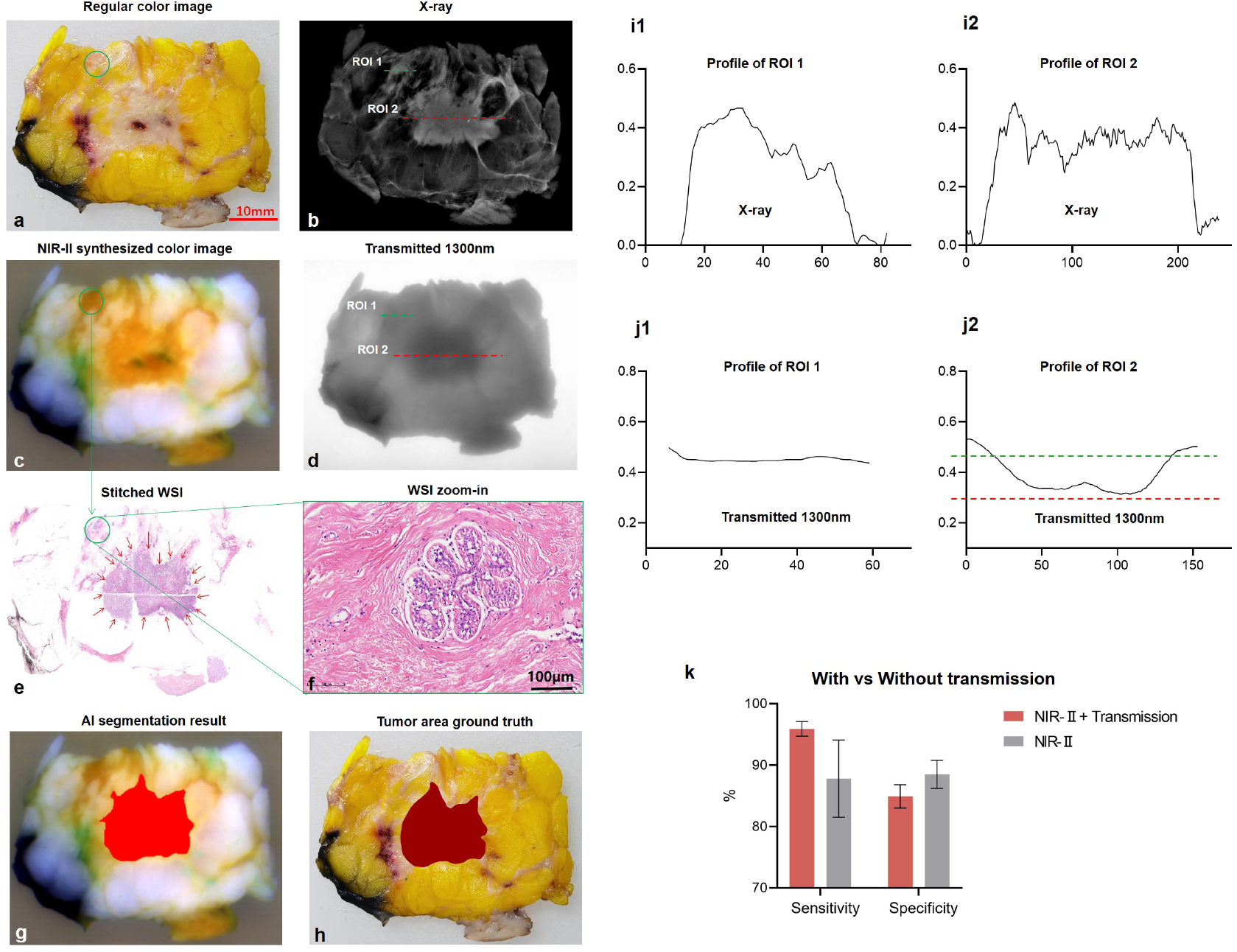
Infrared transmitted light helps differentiate tumor or non-tumor tissues. a. The regular color image of a breast cancer specimen with green circle indicating a normal breast area. b. X-ray image of the specimen. The green and red dash line intensity curves are drawn in i1 and i2 respectively. c. The NIR-II synthesized color image (R: 1050nm, G: 1300nm, B: 1650nm) does not show good contrast between the circled normal area and the tumor area where is circled in dark red in h. d. The 1300nm transmitted image demonstrates the shadow in the tumor area while no shadow in the circled normal area. The green and red dash line intensity curves are drawn in j1 and j2 respectively. e: The tissue in the green circle is confirmed to be dense normal breast tissue in WSI. f: is an enlarged image (HE 20×). g: AI segmentation result. h: Tumor area ground truth. k. statistic results show that incorporating transmitted bands has positive contribution to the segmentation performance of DNMIS.

## Discussion

In this paper, we introduced a multi-spectral imaging system equipped with AI models, DNMIS, to assist pathologists in specimen sampling. We trained an AI algorithm specifically for multispectral breast data to analyze the information-rich multispectral data more completely and rapidly, and obtained objective tumor foci information that can assist the pathologists to judge the scope of the cancer foci, thereby improved the accuracy of the material selection.

Our primary goal was to identify the tumor tissue (including invasive carcinoma and carcinoma in situ). During pathological sampling, in order to ensure that the surgical margin is negative during the operation, the pathologist often adopts the method of vertical incision margin. It is necessary to identify the tumor tissue and measure the distance between the tumor tissue and resection margin to ensure that the closest position of the tumor tissue to the resection margin is taken into consideration, so as to clarify the situation of the resection margin^35,36^. For breast cancer specimens after neoadjuvant therapy, all possible residual tumors need to be identified to avoid inaccurate results of the pathological evaluation due to missed sampling. In particular, if non-pathologic complete response (nPCR) patients are misjudged as PCR, then these patients can lose the opportunity for follow-up treatment, affecting patient survival^37,38^. Therefore, in this study, we developed the DNMIS system, and verified the ability of the system to distinguish tumor tissues from other tissues. The intuitive results showed that whether it is a non-neoadjuvant breast cancer specimen or a breast cancer specimen after neoadjuvant therapy, the NIR-II synthesized color images obtained by the DNMIS system showed clearer tumor boundaries than regular color images and X-ray images. Tumor tissue can be distinguished from the normal breast tissue, adipose tissue, and benign breast lesions, which is helpful for pathologists. In particular, it can identify the fibrotic tumor bed of the residual cancer tissue after neoadjuvant therapy, which is an obvious advantage over X-ray. Although X-rays can identify tumor beds, their ability to identify fibrotic tumor beds and tumor foci is limited. The research results showed that for the 80 samples collected in this study, the sensitivity of AI segmentation based on the multispectral image information of the DNMIS system to identify tumor tissue was as high as 95.9%in sensitivity, which means that 95.9% of the tumor tissues can be detected by DNMIS better than the pathologist’s visual inspection alone (88.4% sensitivity) and with the aid of X-ray images (92.8% sensitivity); AI assisted DNMIS also had a competitive specificity (DNMIS 84.9% vs Pathologist-visual 88.2% vs Pathologist-visual-Xray 83.4%), In particular, for specimens obtained after neoadjuvant treatment, the sensitivity of tumor identification solely by visual observation was only 68.6%, and specificity was 89.7%. The importance of pathological grading of the patient’s tumor bed is underestimated, which may result in the patient losing the opportunity to continue treatment. The sensitivity of AI segmentation results using DNMIS to identify tumor tissue was as high as 90.3%. Therefore, the detection rate of tumor tissue can be greatly improved with the assistance of this system. After neoadjuvant therapy, the tumor bed of breast cancer shows different regression patterns (e.g. non-centripetal regression pattern), making it difficult to distinguish the scattered tumor tissue in the tumor bed with visual inspection. Our study indicates that NIR multispectral imaging technology and AI algorithm can significantly improve the limits of human perception.

In our study, with limited specimen number (80 cases), we found that human experience-based wave selection can help improve the AI segmentation results. For example, removing visible bands from the collected data improves the AI performance. However, if the training data is very plentiful, then the band selection maybe unnecessary as usually more information should be better. Chemical properties of the colored inks that absorb less infrared light. Reflected imaging can face challenge in distinguishing dense fibrous connective tissue from cancerous tissue; hence, NIR transmitted light was added to our experiments. The subsequent results indicated that dense fibrous connective tissue and tumor tissue showed different light transmittance, and the collected NIR transmission images of the specimens were viable alternatives to conventional X-ray images. The X-ray image is intensity based and the only output is a single spectral channel, which provides relatively limited information. Conversely, our method requires no ionizing radiation and has a lower cost compared to the X-ray approach.

As far as the current technology is concerned, our method cannot accurately distinguish between invasive carcinoma and intraductal carcinoma, both of which will be identified as tumor tissue at the same time. However, in terms of the role in guiding specimen sampling, it is not so important to accurately distinguish between the two.

Nonetheless, for future clinical applications, DNMIS still requires several improvements. For now, the AI segmentation results were shown on the monitor to guide the pathologists during specimen sampling. However, a matching error can occur when the pathologists switch views between the monitor and the specimen. One solution maybe to use augmented reality to display segmented tissue margin and cutting scheme onto the specimen surface, so that the pathologists can have the optimal sampling especially for large tumors where multiple slides are needed. For example, we can use a fixed projector to project the tumor outline on the specimen to guide the specimen sampling in real-time and in situ. Another limitation is that the SWIR camera has low spatial resolution (640×512), which may limit the segmentation fineness. The SWIR camera is InGaAs-based and has a wider sensing range but has poor image resolution compared to a regular color camera (Si-based). One solution is to use an imaging lens with a larger magnification and mechanically scan the sample to stitch a high-resolution image. With the maturity of the semiconductor technology, the resolution of SWIR cameras will go higher and higher. For example, Sony has launched the world’s highest resolution SWIR sensor in 2021: IMX990, which has a resolution of 1280×1024.

In spite of these limitations, DNMIS exhibits high sensitivity and specificity in breast tissue classification and can assist pathologists in achieving complete tumor sampling. Given the promise of our initial experiments in ex vivo specimen sampling, the proposed method may also find application in intraoperative cancer margin assessment, which may help in cancer residue dissection, thus avoiding a second surgery for patients.

## Methods

### Preparation of tissue samples and experimental procedures

A total of 80 breast cancer resection specimens were collected, including 16 specimens from patients who received neoadjuvant therapy. All specimens were placed in a sufficient amount of 3.7% neutral formalin solution 30 minutes after being ex vivo, and the fixation time was 12–48 h. The tissue slices were cut into sections with a thickness of 5 mm±1 mm, with an average of 5 mm, including the tumor tissue and surrounding normal tissue of 1-2 cm. Conventional color images were taken from the same angle for all samples, X-ray images were taken by a cabinet X-ray imaging system, and multispectral images were taken by a DNMIS device. The tissue slices were obtained by gross sampling, routine dehydration, paraffin embedding, hematoxylin and eosin (HE) staining, and scanning to obtain several WSI fragments. A self-developed WSI stitching software was used to stitch WSI fragments to restore virtual large slices. The tumor bed range marked by the pathologist using the ASAP marking software on the virtual large slice was used as the “gold standard”. Five pathologists with more than 3 years of work experience marked the contours of the tumor tissue on the conventional color images with reference to the conventional color images and conventional color images + X-ray images, as the results of the traditional sampling method. Subsequently, we compared AI labeling of tumor tissues over the traditional methods.

### Hardware components

The camera of DNMIS is ARTCAM-991SWIR-TEC from ARTRAY Inc., Japan. It contains the Sony IMX991 image sensor, which has a high quantum efficiency in the range of 400–1700 nm and a resolution of 640×512. In comparison, regular Si-based or InGaAs-based sensors can only sense visible or NIR-II. Our camera choice made it possible to capture images covering the visible to the NIR-II wavelength with a single camera. A 12.5 mm broad band lens LM12HC-SW from Kowa Co. Ltd. was used as the main imaging lens. A liquid tunable lens EL-16-40-TC by Optotune Inc. was installed in front of the main lens for autofocus adjustment. For the customized multispectral light source, each reflected illumination wavelength has an output of 15 W power and each transmitted illumination wavelength has an output of 30 W power. We used multi-channel light controllers to adjust the light intensity and switching between different wavelength light emitting diodes (LEDs). A black matt cabinet was used as the outer packaging of the system to block the ambient light. In order to clearly show the core components of the system, this cabinet is not drawn in Fig.1.

### Reflectance correction for reflected images

A whiteboard with 99.9% reflectance was used for reflectance correction. Before image acquisition for each specimen, the whiteboard was put above the translucent frosted acrylic sheet to capture a series of images for reflectance correction. The formula for reflectance correction is as follows:

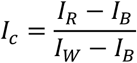

Where *I*_*c*_ represents the corrected image; *I*_*R*_ represents the raw multispectral image; *I*_*B*_ is the black environment image captured when all light sources are turned off; *I*_*W*_ is the whiteboard image captured when the whiteboard is put above the acrylic sheet.

### Anti-glaring correction for reflected images

To remove the glare in the reflected images, we used a cross-polarization technique^39^ to improve the image quality. Accordingly, we installed a broadband wire grid polarizer on the surface of the reflected light sources. To achieve cross polarization, we also installed the same polarizer in front of the main lens, but with a 90° polarization angle difference with the ones on the light sources.

### High dynamic range (HDR) for transmitted images

To eliminate the influence of different specimen thickness on the transmitted light, we introduced the HDR function to process the transmitted images. HDR requires the imaging system to capture images of the sample with different exposure times in order to brighten the details of the dark parts, while keeping the highlights from being overexposed. Here, we used the state of the art HDR algorithm^40^ with five exposures: 1/15s, 1/30s, 1/60s, 1/100s, 1/200s.

### Registration of different image sources

Since the data collected by DNMIS included dual-mode multispectral images, and all datasets included regular color images, X-ray images, and WSI as the gold standard from the same perspective, we needed to register the multi modal data. For each sample, we use the regular color image as the reference image. All other types of images including X-ray, multispectral and WSI images will be registered with the reference image. In the registration process, we manually choose at least four pairs of matching points between the image to be registered and the reference image, and then calculated the affine transformation matrix to preliminarily register the different modal images. For local areas where tissue deformation exists between different type of images, pathologists will reference all image evidences to figure out the deformation and select extra local matching points to correct it. Since the different refractive indices of different illumination wavelengths in DNMIS will lead to inconsistent image magnification, the chromatic aberration problem between multispectral images can be effectively solved by the above registration process.

### AI algorithm details for tissue segmentation

As a classical semantic segmentation network, U-net can effectively extract the feature information from samples in both spatial and spectral dimensions^41^. We used the multispectral images collected by DNMIS and the corresponding ground truth labels as training pairs for supervised training. All samples are divided evenly into 3 folds. During this process, we ensured that the neoadjuvant and non-neoadjuvant samples were evenly distributed in each fold. Before feeding the images to U-net, the images were cut into 208×208 patches, and then we performed data augmentation steps, including random scaling and rotation. The number of epochs for training was 200. The neural networks were implemented and trained using TensorFlow on an NVIDIA GTX 1060 GPU, and the training process took 2 hours. After training, the neural networks with the optimized parameters were used for fast inference almost in real time, which amounted to just 0.8 second for each sample.

## Data Availability

All data produced in the present study are available upon reasonable request to the authors.

## Acknowledgements

This work is supported by the Beijing Fine Inspection Foundation, (Grant No. JJIS2021-011). This study is also supported by Tencent AI Lab, the Fourth Hospital of Hebei Medical University and the West China Hospital, Sichuan University. We thank the pathologists in the Fourth Hospital of Hebei Medical University for collecting the specimens. We thank all engineers in Tencent AI Lab for data processing and analysis of images.

## Author contributions

Y.P.L., J.H.Y, and H. B. conceived of the idea and directed the work. J.L. and L.L.Z. performed the experiments, analyzed the data, wrote and edited the manuscript. H.W. set up the hardware system. Z.Q.B. and C.C.Q. assisted in deep learning network training of the proposed system. M.Z., D.D.H, Z.L.J collected tissue samples and completed the collection of conventional color images, X-ray and specular images. All pathologists participated in sample labeling. All authors wrote and revised the manuscript.

## Additional information

The study was approved by the ethics committee of the Fourth Hospital of Hebei Medical University. Written informed consent was obtained from all of participants according to the study protocols.

## Notes

### Competing Interest Statement

The authors have declared no competing interest.

### Author Declarations

Ethics committee of the Fourth Hospital of Hebei Medical University gave ethical approval for this work.

